# Urban, regional and remote outcomes following TAVR for Queensland patients: a retrospective cohort study

**DOI:** 10.64898/2026.10.22.26366273

**Authors:** Patrick Cameron, Anthony Brazzale, Greg Starmer, Joseph V Moxon

## Abstract

**Background:** Transcatheter aortic valve replacement (TAVR) offers a minimally invasive approach to treat severe aortic stenosis (AS) but can only be performed at centres meeting strict regulatory requirements. It remains unclear whether this requirement restricts TAVR access for patients residing in rural Australia, and whether such people experience poorer outcomes than metropolitan counterparts as a result.

**Methods:** This multicentre retrospective cohort study used Australian Cardiac Outcomes Registry data. All Queensland residents who underwent TAVR between April 2018 and 27 March 2026 were included. Rurality was assigned by postcode using the Modified Monash Model (MMM) and grouped as urban (MMM1), regional (MMM2–4), or remote (MMM5–7). The primary outcome was 12-month mortality and was investigated using Kaplan-Meier and Cox proportional hazards analyses.

**Findings:** Among 6813 patients, 4246 (62%) were urban, 2092 (31%) regional, and 475 (7%) remote. Twelve-month all-cause mortality did not differ by rurality (urban: 211 [5.0%]; regional: 103 [4.9%]; remote: 23 [4.8%]; χ²=0.991, log-rank P=0.929), a finding confirmed in multivariable analysis. At the time of TAVR, regional and remote patients were younger, more comorbid, had more severe AS, and lower STS scores than urban counterparts. Clinical frailty score, non-transfemoral access, STS score, atrial fibrillation or flutter, and moderate–severe chronic lung disease independently predicted 12-month mortality.

**Interpretation:** TAVR outcomes were comparable between rurality groups, suggesting equitable care within Queensland’s centralised TAVR model. However, registry data only captures patients who underwent TAVR and may miss older rural patients facing barriers to AS diagnosis, referral, or procedural workup.

**Funding:** There was no funding or sponsor involvement.

**Research in context:** *Evidence before this study:* We searched Medline (Ovid) for publications examining the association between rurality and outcomes after transcatheter aortic valve replacement (TAVR). We searched from database inception to September 2026 using the terms “TAVR” OR “transcatheter aortic valve replacement” OR “transcatheter aortic valve implantation” OR “TAVI”, AND “rural*”. We identified two key studies of rurality and TAVR outcomes, one from the USA and one from Australia. Both reported no difference in mortality between rural and urban patients. However, both were single-centre studies with small samples (413 and 142 patients, respectively) and neither used time-to-event analysis. The US study followed patients for only 6 months and used descriptive analysis alone. The Australian study included urban and regional patients but not remote patients.

*Added value of this study:* In 6813 patients from a national procedural registry, 12-month all-cause mortality after TAVR did not differ between urban, regional, and remote Queensland residents (5.0%, 4.9%, and 4.8%, respectively), including after multivariable adjustment and in sensitivity analyses. This was despite regional and remote patients being younger, having more comorbidity, lower STS scores, and having more severe AS at the time of the procedure. To our knowledge, this is the largest study of TAVR outcomes by rurality and the first in Australia to include remote patients. Remote residents and First Nations people were underrepresented among TAVR recipients compared with the Queensland population.

*Implications of all the available evidence:* Among people who reach TAVR, a centralised delivery model appears to provide comparable short-term outcomes regardless of where patients live. However, ACOR data only captures patients who received the procedure, and the younger age of rural patients and underrepresentation of remote patients suggests that barriers may exist earlier in the pathway, at diagnosis, referral, or work-up. Future research linking diagnostic and referral data to procedural data is needed to assess access, and the findings should inform planning of non-metropolitan AS services.

## Introduction

Aortic stenosis (AS) is the most common valvular heart disease in the developed world and affects approximately 12% of people over the age of 75 years.^1,2^ Aortic valve replacement is the only effective treatment for severe AS, without which, 2– and 5-year survival rates are 50% and 20% respectively.^3^ Surgical aortic valve replacement (SAVR) is the traditional approach to treat severe AS, but is highly invasive meaning that up to 30% of AS patients may be ineligible due to advanced age, left ventricular dysfunction or presence of multiple coexisting conditions.^4^ The development of transcatheter aortic valve replacement (TAVR), whereby the diseased aortic valve is replaced with a bioprosthetic implant via the femoral artery, offers an alternative approach to treat patients otherwise ineligible for SAVR.^4^ TAVR is now widely adopted and the use of this procedure is expected to expand, with studies forecasting 27,000 new TAVR patients annually across the Americas and Europe.^1^

Access to TAVR in Australia remains concentrated in major centres as regulatory requirements mandate minimum institutional and operator procedural volumes and on-site cardiothoracic surgery.^5^ This has particular implications for rural Australians, who experience reduced access to healthcare, fewer general practitioner visits, and limited availability of cardiac specialists, often requiring long-distance transfer to tertiary referral centres for definitive management.^6–8^ Together, these factors may contribute to later presentation with more severe AS, which has been associated with worse outcomes following TAVR.^9–11^ In addition, greater comorbidity burden and reduced access to post-procedural cardiac rehabilitation for rural patients may further influence recovery and longer-term outcomes.^7,12–15^ However, evidence comparing TAVR outcomes between rural and metropolitan patients remains limited, motivating this study.

The Australian state of Queensland provides a relevant context in which to address this evidence gap. The state has the highest proportion of residents living outside of major cities of any Australian state, with 35% of the population based in regional areas.^16^ Furthermore, 2% of the population live in remote (MMM6) or very remote (MMM7) areas, where access to specialised cardiac services including TAVR is limited.^16^ This population distribution creates unique challenges for delivering equitable TAVR services, making Queensland an ideal setting to explore differences in TAVR outcomes between regional and metropolitan patients in Australia. Accordingly, this study aims to determine whether outcomes following TAVR differ for patients residing in regional and remote compared to urban Queensland.

## Methods

### Study design

This multicentre retrospective, observational cohort study used data from the Australian Cardiac Outcomes Registry (ACOR), a nationwide cardiac care registry with mandated submission of procedural data under the Australian Medicare Benefits Schedule, ensuring standardised national data capture.^17^ We collected data for all patients who resided in Queensland at the time of their TAVR, between April 2018 (first ACOR entry) and the 27^th^ of March 2026. Data for 6,847 Queensland TAVR patients was collected. Nine cases were excluded due to missing Modified Monash Model (MMM) classification and 25 due to aborted procedure, leaving a final cohort of 6,813 patients for analysis (supplementary Figure 1). The study protocol was approved by the local Human Research Ethics Committee and James Cook University (HREC/2025/QCH/117308), and a waiver of consent to access data was granted. This study was reported according to the STROBE statement for cohort studies (supplementary Figure 2).

### Patients

All Queensland residents with an MMM classification who underwent a TAVR recorded in the ACOR database within the sampling period were eligible. MMM incorporates the Australian Statistical Geography Standard-Remoteness Areas (ASGS-RA), road distance between towns and population size of localities to classify areas from major cities (MMM1) to very remote communities (MMM7).^18^ Patient’s residential postcodes were used to assign a MMM rurality classification.^19^ When a postcode included multiple MMM classifications, the least rural classification was selected as it was assumed this would contain the most people.^20^ For the analysis, patients were grouped as urban (MMM1), regional (MMM2-4) or remote (MMM5-7).

### Outcomes

The primary outcome was 12-month all-cause mortality. Secondary outcomes were; i) post-procedure length of hospital stay; ii) in-hospital complications (cardiac complication, cerebral complication, vascular complication, bleeding complication, device complication, and renal complication); iii) 30-day and 12-month adverse events (mortality, rehospitalisation for heart failure, stroke, major vascular complication, permanent pacemaker insertion [PPM], bleeding complication, cardiac complication, renal failure requiring dialysis, myocardial infarction [MI], and major adverse cardiac and cerebrovascular events [MACCE]); iv) postprocedural, 30 day, and 12 month echocardiographic parameters (aortic valve [AV] area improvement, AV mean gradient improvement, left ventricular ejection fraction [LVEF] improvement, and paravalvular regurgitation); v) variation in the timing of attending mandated 30 day and 12 month follow up appointments. To allow for natural variation in registry follow-up, ACOR defines 30-day outcomes as those occurring up to 42 days and 12-month outcomes as those occurring up to 14 months (426 days). For the primary mortality analysis, time-to-event was therefore censored at 426 days after TAVR in line with ACOR data definitions.

#### Outcome and missing data

To account for inconsistencies in the dataset, where a valid time-to-death value was available within the ACOR-defined 12-month follow-up window of 426 days, but the “12-month mortality” variable was missing, the “12-month mortality” variable was recoded as a death event. Conversely, where a death event was recorded in the “12-month mortality” variable, but no time-to-death value was available, “time-to-death within 426 days” and “time-to-death within 365 days” variables were coded as missing because the event timing could not be verified. No attempt to impute other missing variables was made and missing datapoints are reported for transparency.

### Statistical analyses

Analyses were conducted using Statistical Package for the Social Sciences (SPSS) version 31 (IBM, Armonk, New York, USA) and R version 4.6.1 (R Foundation for Statistical Computing, Vienna, Austria).

#### Descriptive statistics

Categorical variables are presented as numbers and percentages and were compared using χ2 test or Fisher’s exact test. Continuous variables are presented as median and interquartile ranges and compared using the Kruskal-Wallis or Mann-Whitney U test as they were not normally distributed on Shapiro-Wilkes testing. Given the large number of cases, the Benjamini-Hochberg method was applied to cross-sectional analysis to reduce the risk of type 1 error, accepting q-values <0.05 as statistically significant.

#### Cox regression

The association of rurality with the primary outcome (12-month mortality) was assessed using Kaplan-Meier analysis with log-rank testing to compare between groups and Cox proportional hazards models. Unadjusted cox-regression analyses were initially conducted to identify characteristics potentially influencing the risk of experiencing the outcome (Supplementary Table 1). Multivariable cox analyses were then conducted incorporating covariates selected using a hybrid approach. *A priori* covariates were selected if they were established predictors of cardiovascular or TAVR mortality and included age, gender, BMI, coronary artery disease, diabetes mellitus, chronic kidney disease (CKD), and prior MI.^12,13,15,21^ Further covariates were included if they showed a potential association with 12-month mortality risk following univariate Cox regression (accepting p<0.05) and included clinical frailty score ≥5, Society of Thoracic Surgeons (STS) score, atrial fibrillation (AF) or flutter, previous IECD (implanted cardiac electrical device), peripheral artery disease, congestive heart failure, New York Heart Association (NYHA) class III-IV, moderate-severe chronic lung disease, AV area, mean AV gradient, LVEF, previous balloon aortic valvuloplasty (BAV), and vascular access site.

Collinearity was defined as a Pearson correlation coefficient exceeding 0.7, and the variable which was considered to have less clinical significance was excluded from multivariable modelling (Supplementary Figure 3). All presented models conformed to the proportional hazards assumptions which were assessed using log-minus log plots for categorical variables and partial residuals for continuous variables. Given 337 events, the number of covariates was limited to maintain an events-per-variable ratio of at least 10:1.

#### Sensitivity analysis

Given that the primary analysis for 12-month mortality was censored at 426 days (in line with ACOR definitions), a sensitivity analysis censored at 365 days was performed to assess whether delayed follow-up influenced outcomes. A second sensitivity analysis was performed with rurality dichotomised as urban (MMM1) versus non-urban (MMM2–7). This reflected the concentration of TAVR services in major metropolitan centres, with most areas outside MMM1 having limited direct access to TAVR services, except for selected MMM2 locations. For all Cox Models, p-values <0.05 were considered significant.

## Results

### Study population and baseline characteristics

Of the 6813 patients included in this study, 4246 (62%) resided in urban areas (MMM1), 2092 (31%) were resident in regional areas (MMM2-4) and 475 (7%) resided in remote areas (MMM5-7). Age at the time of TAVR decreased as rurality increased while the proportion of patients who identified as First Nations increased as rurality increased (Table 1). The proportion of patients who were male was highest in the remote and lowest in the regional group (Table 1). BMI, diabetes, moderate-severe chronic lung disease, smoking, mean AV gradient, peak AV velocity, and use of balloon-expanding valves all increased with increasing rurality (Table 1). In contrast, STS score, DP index and use of cerebral protection devices decreased with increasing rurality (Table 1). Prior MI, congestive heart failure, NYHA status III or IV and smaller aortic valve area were more common in the regional and remote groups than urban, whilst transfemoral access route was lower in regional compared to urban and remote patients (Table 1).

**Table 1.** Baseline demographics, clinical, echocardiographic and procedural characteristics by MMM group.

| Variable | Urban<br>(n=4246) | Regional<br>(n=2092) | Remote<br>(n=475) | p<br>value | q<br>value |
| --- | --- | --- | --- | --- | --- |
| **Classification** | MMM1 | MMM2-4 | MMM5-7 |  |  |
| **<u>Demographics</u>** |  |  |  |  |  |
| **Age (years)** | 82 (77-86) [2] | 81 (76-85) [4] | 80 (76-84) | <0.001 | 0.003 |
| **Male gender, n (%)** | 2594 (61.1%)<br>[2] | 1272 (60.8%)<br>[1] | 323 (68.0%)<br>[1] | 0.009 | 0.019 |
| **First Nations, n (%)** | 17 (0.4%) [223] | 19 (0.9%) [87] | 8 (1.7%) [20] | <0.001 | 0.003 |
| **<u>Clinical characteristics</u>** |  |  |  |  |  |
| **Body mass index (kg/m<sup>2</sup>)** | 27.68 (24.44-31.60) [12] | 28.38 (25.25-32.41) | 28.91 (25.36-32.79) | <0.001 | 0.003 |
| **Clinical frailty score ≥5, n (%)** | 1008 (23.7%)<br>[272] | 532 (25.4%)<br>[92] | 119 (25.1%)<br>[20] | 0.579 | 0.579 |
| **STS score (%)** | 4.20 (2.50-7.10) [413] | 3.99 (2.44-6.80) [371] | 3.56 (2.29-6.00) [69] | 0.002 | 0.005 |
| **Coronary artery disease, n (%)** | 2583 (60.8%)<br>[13] | 1338 (64.0%)<br>[1] | 292 (61.5%) | 0.071 | 0.108 |
| **Previous myocardial infarction, n (%)** | 449 (10.6%)<br>[12] | 309 (14.8%)<br>[7] | 56 (11.8%) | <0.001 | 0.003 |
| **Previous PCI, n (%)** | 957 (22.5%) [5] | 474 (22.7%)<br>[2] | 97 (20.4%) | 0.544 | 0.562 |
| **Previous cardiovascular surgery, n (%)** | 1053 (24.8%)<br>[8] | 472 (22.6%)<br>[2] | 119 (25.1%)<br>[3] | 0.122 | 0.178 |
| **Previous balloon aortic valvuloplasty, n (%)** | 141 (3.3%) [5] | 61 (2.9%) [2] | 11 (2.3%) [1] | 0.394 | 0.420 |
| **Peripheral artery disease, n (%)** | 718 (16.9%) [9] | 341 (16.3%)<br>[1] | 60 (12.6%)<br>[1] | 0.056 | 0.093 |
| **Previous stroke or TIA, n (%)** | 487 (11.5%) [6] | 266 (12.7%)<br>[6] | 49 (10.3%)<br>[1] | 0.203 | 0.25 |
| **Atrial fibrillation or flutter, n (%)** | 1348 (31.7%)<br>[7] | 655 (31.3%)<br>[3] | 167 (35.2%)<br>[1] | 0.256 | 0.293 |
| **Classification** | MMM1 | MMM2-4 | MMM5-7 |  |  |
| **Previous implanted cardiac electrical device, n (%)** | 699 (16.5%) [8] | 313 (15.0%) | 68 (14.3%) | 0.184 | 0.237 |
| **Congestive heart failure, n (%)** | 1049 (24.7%) [91] | 655 (31.3%) [55] | 137 (28.8%) [6] | <0.001 | 0.003 |
| **NYHA class III or IV, n (%)** | 2294 (54.0%) [78] | 1200 (57.4%) [42] | 271 (57.1%) [6] | 0.026 | 0.049 |
| **Hypertension, n (%)** | 3136 (73.9%) [2] | 1589 (76.0%) [2] | 355 (74.7%) | 0.185 | 0.237 |
| **Diabetes, n (%)** | 1069 (25.2%) [3] | 613 (29.3%) [1] | 144 (30.3%) | <0.001 | 0.003 |
| **Chronic kidney disease (eGFR < 60), n (%)** | 1506 (35.5%) [90] | 761 (36.4%) [57] | 157 (33.1%) [8] | 0.290 | 0.320 |
| **Moderate–severe chronic lung disease, n (%)** | 569 (13.4%) [41] | 318 (15.2%) [21] | 81 (17.1%) [5] | 0.028 | 0.0498 |
| **Current or recent smoker (<1 year), n (%)** | 115 (2.7%) [21] | 77 (3.7%) [7] | 27 (5.7%) [1] | <0.001 | 0.003 |
| **<u>Echocardiographic parameters (pre-procedure)</u>** |  |  |  |  |  |
| **Aortic valve area (cm<sup>2</sup>)** | 0.90 (0.70-1.00) [131] | 0.80 (0.70-0.97) [57] | 0.81 (0.70-1.00) [10] | <0.001 | 0.003 |
| **Mean aortic valve gradient (mmHg)** | 41 (33-49) [47] | 42 (36-51) [26] | 43 (36-51) [5] | <0.001 | 0.003 |
| **Peak aortic valve velocity (m/s)** | 4.10 (3.70-4.40) [117] | 4.18 (3.90-4.50) [64] | 4.20 (3.90-4.50) [11] | <0.001 | 0.003 |
| **DP index** | 0.23 (0.20-0.27) [495] | 0.22 (0.19-0.26) [224] | 0.22 (0.19-0.25) [49] | <0.001 | 0.003 |
| **Left ventricular ejection fraction (%)** | 61 (55-65) [26] | 61 (54-66) [21] | 60 (54-65) [4] | 0.144 | 0.200 |
| **Bicuspid aortic valve, n (%)** | 231 (5.4%) [105] | 101 (4.8%) [65] | 18 (3.8%) [14] | 0.242 | 0.287 |
| **<u>Procedural characteristics</u>** |  |  |  |  |  |
| **Classification** | MMM1 | MMM2-4 | MMM5-7 |  |  |
| **Procedure indication, n (%)** | [15] | [2] | [1] | 0.058 | 0.093 |
| – **Predominant AS** | 3709 (87.4%) | 1853 (88.6%) | 408 (85.9%) |  |  |
| – **Predominant AR** | 28 (0.7%) | 16 (0.8%) | 4 (0.8%) |  |  |
| – **Mixed AS/AR** | 147 (3.5%) | 52 (2.5%) | 9 (1.9%) |  |  |
| – **Bioprosthetic valve failure** | 347 (8.2%) | 169 (8.1%) | 53 (11.2%) |  |  |
| **Transfemoral access route, n (%)** | 4068 (95.8%)<br>[3] | 1974 (94.4%) | 455 (95.8%) | 0.023 | 0.046 |
| **Cerebral protection device used, n (%)** | 321 (7.6%)<br>[653] | 109 (5.2%)<br>[239] | 21 (4.4%)<br>[58] | <0.001 | 0.003 |
| **Balloon-expanding valve, n (%)** | 2195 (51.7%)<br>[14] | 1245 (59.5%)<br>[10] | 284 (59.8%)<br>[2] | <0.001 | 0.003 |

- Continuous variables are reported as median (interquartile range); categorical variables are reported as n (% of total cohort). - Values in square brackets indicate missing values. - *P* values were calculated using Kruskal–Wallis tests for continuous variables and $\chi^2$ tests for categorical variables. - *Q* values were calculated using the Benjamini–Hochberg method. - Abbreviations: AR, aortic regurgitation; AS, aortic stenosis; AV, aortic valve; BMI, body mass index; eGFR, estimated glomerular filtration rate; LOS, length of stay; MMM, Modified Monash Model; NYHA, New York Heart Association; PCI, percutaneous coronary intervention; STS, Society of Thoracic Surgeons; TIA, transient ischaemic attack.

### Twelve-month mortality

There was no significant difference in 12-month mortality between rurality groups (urban: 211 [5.0%]; regional: 103 [4.9%]; remote: 23 [4.8%]; χ²=0.991, log rank P=0.929) (Table 2, Figure 1, Supplementary Table 2). Unadjusted and multivariable cox proportional hazard analyses confirmed the lack of association between rurality and 12-month mortality (HR 1.051, (95% CI 0.805–1.373), and 1.039, (95% CI 0.644–1.676) for regional and remote patients respectively compared to urban patients) (Table 3, Supplementary Table 1). In the multivariable model, clinical frailty score ≥5, higher STS score, AF or flutter, moderate-severe chronic lung disease and alternative access were independently associated with increased hazard of death (Table 3).

**Figure 1:**
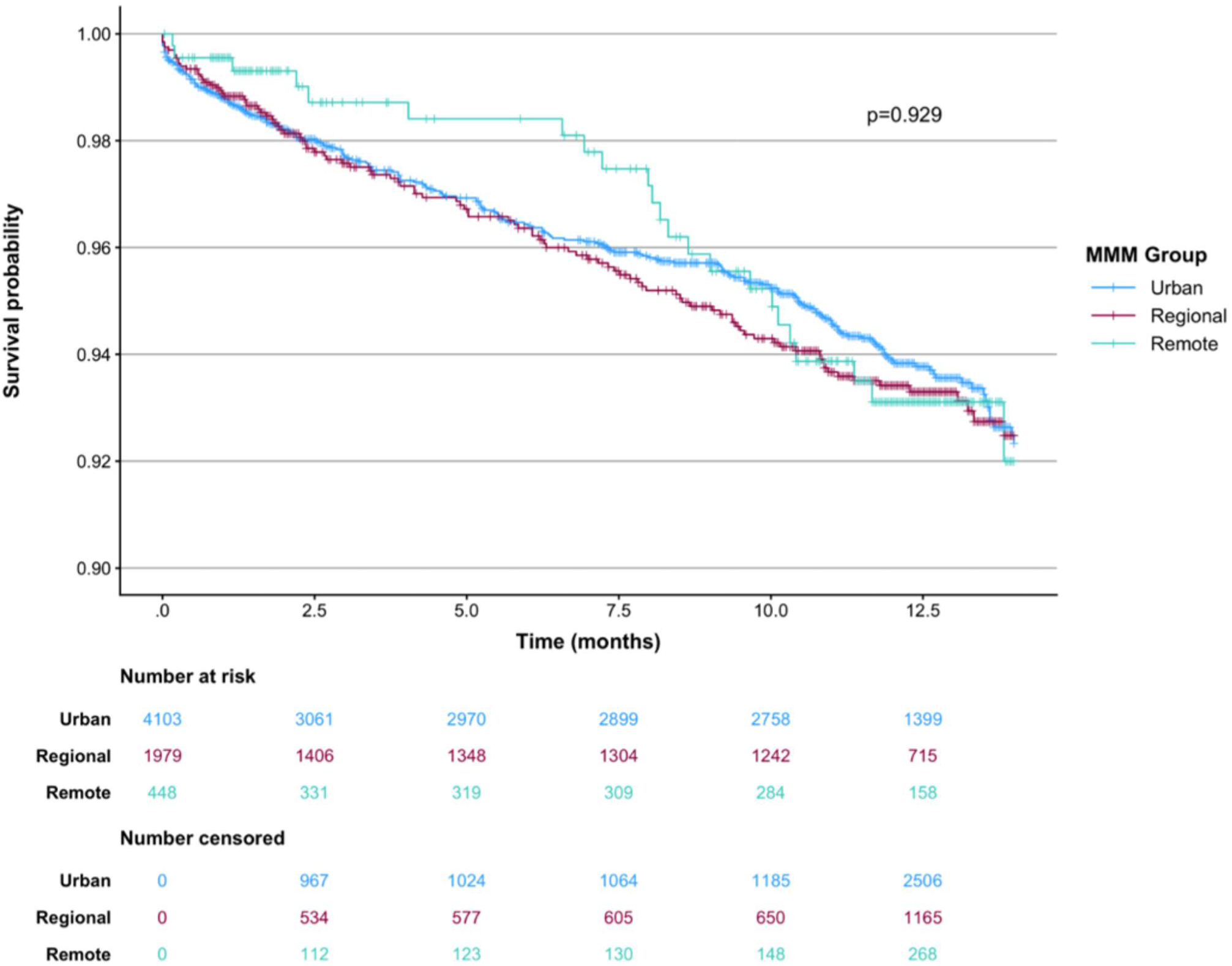
Kaplan-Meier Curve for 12-Month All-Cause mortality stratified by MMM group.

**Table 2.** Outcomes by MMM group.

| Variable | Urban<br>(n=4246) | Regional<br>(n=2092) | Remote<br>(n=475) | p value | q value |
| --- | --- | --- | --- | --- | --- |
| Classification | MMM1 | MMM2–4 | MMM5–7 |  |  |
| **In hospital outcomes** |  |  |  |  |  |
| **Clinical outcomes** |  |  |  |  |  |
| Post-procedure hospital LOS (days) | 2 (2-4) [16] | 3 (2-4) [3] | 3 (2-5) [1] | <0.001 | 0.006 |
| Cardiac complication, n (%) | 469 (11.0%) | 248 (11.9%) | 67 (14.1%) | 0.117 | 0.296 |
| Cerebral complication, n (%) | 62 (1.5%) | 40 (1.9%) | 3 (0.6%) | 0.097 | 0.261 |
| Vascular complication, n (%) | 99 (2.3%) | 47 (2.2%) | 9 (1.9%) | 0.828 | 0.962 |
| Bleeding complication, n (%) | 69 (1.6%) | 34 (1.6%) | 8 (1.7%) | 0.995 | 0.995 |
| Device complication, n (%) | 31 (0.7%) | 21 (1.0%) | 5 (1.1%) | 0.460 | 0.706 |
| Renal complication, n (%) | 47 (1.1%) | 29 (1.4%) | 5 (1.1%) | 0.603 | 0.847 |
| **Echocardiographic outcomes** |  |  |  |  |  |
| AV area improvement (cm<sup>2</sup>) | 1.20 (0.80-1.60) [615] | 1.30 (0.9-1.7) [231] | 1.27 (0.90-1.80) [53] | <0.001 | 0.006 |
| AV mean gradient improvement (mmHg) | 31 (22-39) [177] | 33 (25-41) [78] | 33 (24-41) [15] | <0.001 | 0.006 |
| LVEF improvement (%) | 1 (-3-6) [264] | 1 (-4-5) [112] | 0 (-5-5) [19] | <0.001 | 0.006 |
| Paravalvular regurgitation ≥2+, n (%) | 172 (4.1%) [213] | 61 (2.9%) [94] | 13 (2.7%) [17] | 0.037 | 0.152 |
| **30-day Outcomes** |  |  |  |  |  |
| **Clinical outcomes** |  |  |  |  |  |
| 30-day mortality, n (%) | 60 (1.4%) [163] | 26 (1.2%) [121] | 3 (0.6%) [29] | 0.378 | 0.650 |
| 30-day follow-up time (days) | 37 (31-48) [185] | 41 (33-54) [133] | 39 (32-52) [28] | <0.001 | 0.006 |
| 30-day rehospitalisation for heart failure, n (%) | 86 (2.0%) [283] | 37 (1.8%) [182] | 7 (1.5%) [34] | 0.647 | 0.847 |
| 30-day stroke, n (%) | 63 (1.5%) [427] | 39 (1.9%) [252] | 4 (0.8%) [50] | 0.191 | 0.411 |
| 30-day major vascular complication, n (%) | 40 (0.9%) [432] | 15 (0.7%) [254] | 2 (0.4%) [50] | 0.407 | 0.673 |
| 30-day PPM, n (%) | 406 (9.6%) [437] | 206 (9.8%) [244] | 53 (11.2%) [44] | 0.548 | 0.813 |
| 30-day major bleeding complication, n (%) | 13 (0.3%) [434] | 6 (0.3%) [257] | 1 (0.2%) [49] | 0.936 | 0.995 |
| **30-day cardiac complication, n (%)** | 99 (2.3%)<br>[412] | 55 (2.6%)<br>[248] | 17 (3.6%)<br>[47] | 0.217 | 0.424 |
| **30-day renal failure requiring dialysis, n (%)** | 11 (0.3%)<br>[431] | 11 (0.5%)<br>[256] | 2 (0.4%)<br>[50] | 0.211 | 0.424 |
| **30-day MI, n (%)** | 8 (0.2%)<br>[433] | 4 (0.2%)<br>[260] | 1 (0.2%)<br>[49] | 0.993 | 0.995 |
| **30-day MACCE, n (%)** | 122 (2.9%)<br>[376] | 61 (2.9%)<br>[238] | 7 (1.5%)<br>[47] | 0.190 | 0.411 |
| **Echocardiographic outcomes** |  |  |  |  |  |
| **30-day AV area improvement (cm<sup>2</sup>)** | 1.01 (0.70-1.40)<br>[1046] | 1.08 (0.70-1.46) [543] | 1.10 (0.70-1.51) [122] | 0.093 | 0.261 |
| **30-day AV mean gradient improvement (mmHg)** | 30 (21-38)<br>[743] | 32 (24-39)<br>[382] | 32 (24-39)<br>[85] | <0.001 | 0.006 |
| **30-day LVEF improvement (%)** | 0 (-5-5)<br>[772] | 0 (-5-5)<br>[391] | 0 (-5-5)<br>[80] | 0.081 | 0.261 |
| **30-day paravalvular regurgitation ≥2+, n (%)** | 215 (5.1%)<br>[827] | 72 (3.4%)<br>[395] | 18 (3.8%)<br>[91] | 0.008 | 0.043 |
| **12-month outcomes** |  |  |  |  |  |
| **Clinical outcomes** |  |  |  |  |  |
| **Mortality within 426 days, n (%)** | 211 (5.0%)<br>[24] | 103 (4.9%)<br>[11] | 23 (4.8%)<br>[3] | 0.991 | 0.995 |
| **Mortality within 365 days, n (%)** | 196 (4.6%)<br>[24] | 98 (4.7%)<br>[11] | 22 (4.6%)<br>[3] | 0.993 | 0.995 |
| **12-month mortality, n (%)** | 235 (5.5%)<br>[1420] | 114 (5.4%)<br>[825] | 26 (5.5%)<br>[180] | 0.760 | 0.932 |
| **12-month follow-up time (days)** | 378 (360-415) [1210] | 385 (362-425) [707] | 379 (354-425) [151] | 0.032 | 0.152 |
| **12-month rehospitalisation for heart failure, n (%)** | 76 (1.8%)<br>[1555] | 37 (1.8%)<br>[891] | 12 (2.5%)<br>[185] | 0.448 | 0.706 |
| **12-month stroke, n (%)** | 73 (1.7%)<br>[1746] | 37 (1.8%)<br>[977] | 6 (1.3%)<br>[216] | 0.650 | 0.847 |
| **12-month major vascular complication, n (%)** | 41 (1.0%)<br>[1765] | 14 (0.7%)<br>[990] | 3 (0.6%)<br>[215] | 0.611 | 0.847 |
| **12-month PPM, n (%)** | 373 (8.8%)<br>[1680] | 178 (8.5%)<br>[935] | 42 (8.8%)<br>[203] | 0.765 | 0.932 |
| **12-month major bleeding complication, n (%)** | 13 (0.3%)<br>[1776] | 5 (0.2%)<br>[992] | 1 (0.2%)<br>[216] | 0.929 | 0.995 |
| **12-month cardiac complication, n (%)** | 128 (3.0%)<br>[1750] | 72 (3.4%)<br>[968] | 19 (4.0%)<br>[213] | 0.154 | 0.368 |
| **12-month renal failure requiring dialysis, n (%)** | 10 (0.2%)<br>[1774] | 9 (0.4%)<br>[989] | 2 (0.4%)<br>[215] | 0.270 | 0.505 |
| **12-month MI, n (%)** | 16 (0.4%)<br>[1777] | 9 (0.4%)<br>[992] | 0 (0%)<br>[218] | 0.341 | 0.611 |
| **12-month MACCE, n (%)** | 294 (6.9%)<br>[1537] | 139 (6.6%)<br>[883] | 29 (6.1%)<br>[194] | 0.780 | 0.932 |
| **Echocardiographic outcomes** |  |  |  |  |  |
| **12-month AV area improvement (cm<sup>2</sup>)** | 1.00 (0.60-1.30)<br>[2169] | 1.00 (0.62-1.40)<br>[1215] | 1.00 (0.70-1.40) [277] | 0.097 | 0.261 |
| **12-month AV mean gradient improvement (mmHg)** | 30 (20-38)<br>[1943] | 31 (24-38)<br>[1131] | 32 (22-39)<br>[251] | <0.001 | 0.006 |
| **12-month LVEF improvement (%)** | 0 (-5-5)<br>[1925] | 0 (-5-7)<br>[1107] | 1 (-4-8)<br>[251] | 0.039 | 0.152 |
| **12-month paravalvular regurgitation ≥2+, n (%)** | 164 (3.9%)<br>[1967] | 52 (2.5%)<br>[1120] | 11 (2.3%)<br>[252] | 0.091 | 0.261 |

- Continuous variables are reported as median (interquartile range); categorical variables are reported as n (% of total cohort). - Values in square brackets indicate missing values. - *P* values were calculated using Kruskal–Wallis tests for continuous variables and $\chi^2$ tests for categorical variables. - *Q* values were calculated using the Benjamini–Hochberg method. - Abbreviations: AV, aortic valve; FUP, follow-up; HF, heart failure; LOS, length of stay; LVEF, left ventricular ejection fraction; MACCE, major adverse cardiac and cerebrovascular events; MI, myocardial infarction; MMM, Modified Monash Model; PPM, permanent pacemaker. - Death within 365 days was defined using a confirmed time-to-death value occurring within 365 days after TAVR - Death within 426 days was defined using a confirmed time-to-death value occurring within the ACOR-defined 12-month follow-up window of 426 days - Where “12-month mortality” was recorded but no time-to-death value was available, the observation was coded as missing

**Table 3.**
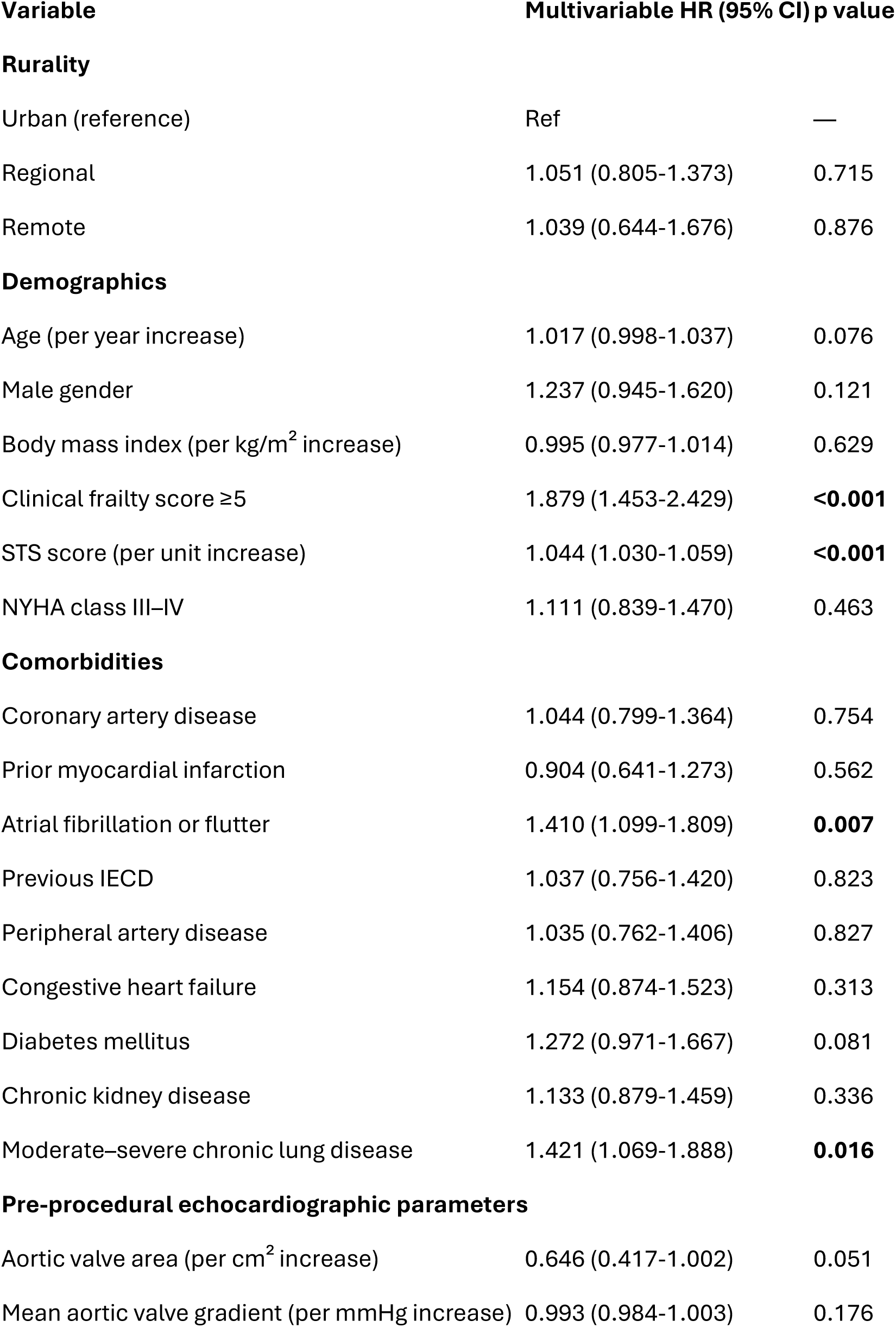
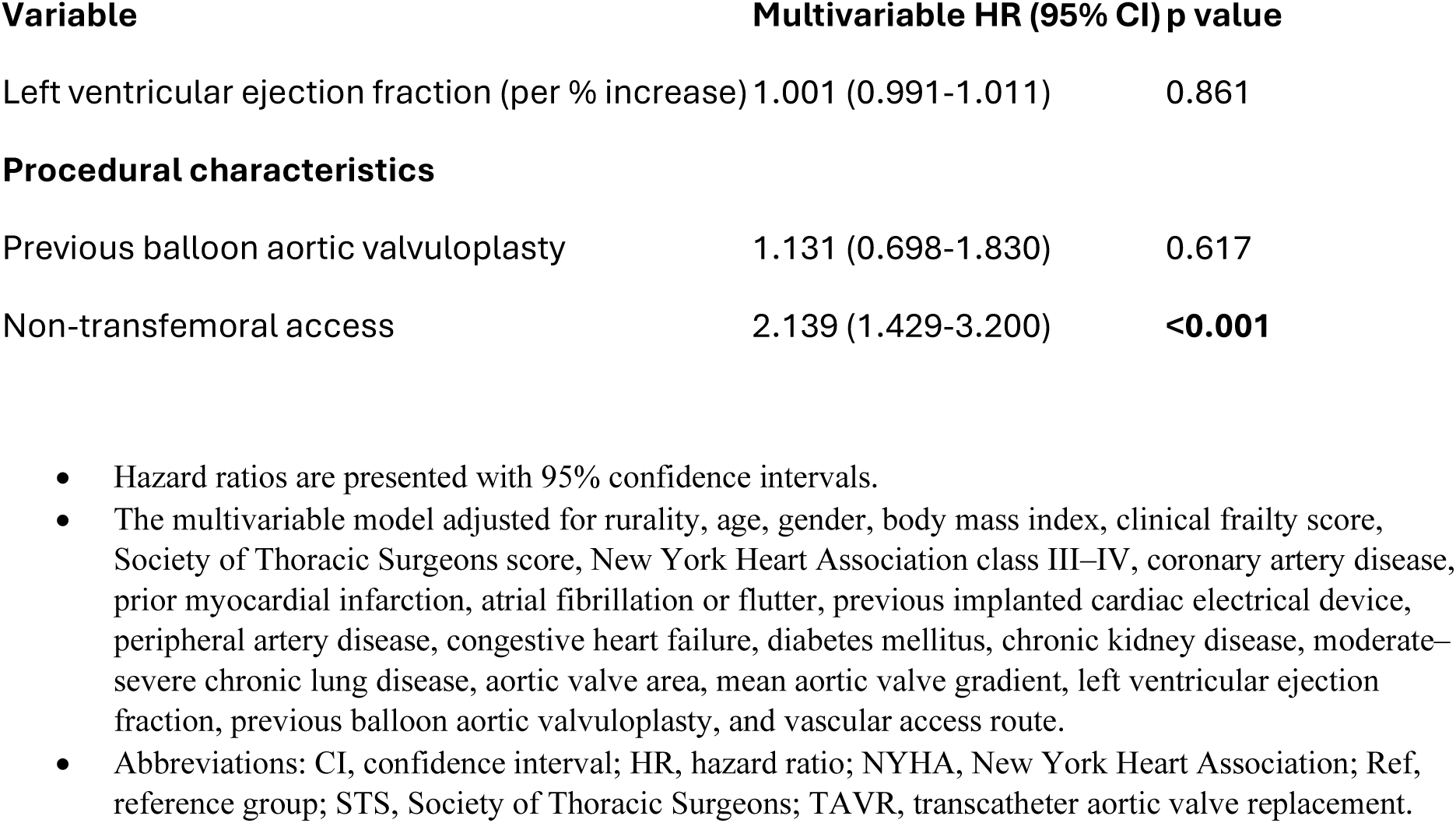
Multivariable Cox proportional hazards regression analysis for 12-month mortality following TAVR.

### Secondary outcomes

The median length of stay in hospital post procedure was one day longer for regional and remote patients (P<0.001, q=0.006), however, an improvement in AV area (P<0.001, q=0.006), AV mean gradient (P<0.001, q=0.006) and LVEF (P<0.001, q=0.006) post procedure were observed in regional and remote patients compared to urban (Table 2). The greater AV mean-gradient improvements for regional and remote patients compared to urban persisted at 30-days (P<0.001, q=0.006) and 12 months (P<0.001, q=0.006), while paravalvular regurgitation ≥2+ was more frequent in urban patients at 30 days (P=0.008, q=0.043) (Table 2). Urban patients had shorter median 30-day follow-up times (P<0.001, q=0.006) (Table 2).

### Sensitivity analyses

Sensitivity analyses restricting follow-up to 365 days upheld the lack of association between rurality and 12-month mortality (HR 1.069, (95% CI 0.812–1.408), and 1.037, (95% CI 0.634– 1.695) for regional and remote patients respectively compared to urban patients) (Supplementary Table 3). A second sensitivity analysis dichotomised rurality as urban (MMM1) versus non-urban (MMM2–7); this also showed no significant association between rurality and 12-month mortality (HR 1.049, 95% CI 0.817–1.347) (Supplementary Table 4).

## Discussion

This multicentre study of 6813 Queensland patients undergoing TAVR found no significant difference in 12-month all-cause mortality between urban, regional, and remote patients, a finding that was confirmed in multivariable and sensitivity analyses. This was observed despite regional and remote patients being younger, having a greater comorbidity burden and more severe AS at the time of procedure, challenging prior evidence linking greater disease severity and comorbidity burden to adverse TAVR outcomes.^9–13^ Similarly, most secondary outcomes were comparable across rurality groups immediately after the procedure, at 30 days and at 12 months, although regional and remote patients had a one-day longer post-procedural length of stay and minor differences in echocardiographic parameters. In multivariable analysis, clinical frailty score ≥ 5, STS score, AF or flutter, moderate-severe chronic lung disease, and non-transfemoral access were independent predictors of 12-month mortality.

Compared with MMM population statistics for Queensland, urban patients were proportionately represented among TAVR recipients (62% in our dataset vs. 63% in the population), whereas regional patients were overrepresented (31% vs. 27%) and remote patients were underrepresented (7% vs. 10%).^20^ The inter-group variation in baseline comorbidities aligns with Australian Institute of Health and Welfare data showing rural populations to be more comorbid,^7^ which may contribute to earlier disease onset and hence the younger median age we observed in these groups. However, the combination of younger rural patients and underrepresentation of remote patients in this dataset may indicate an access disparity, whereby older rural patients are less able or willing to undertake the travel required to access TAVR in a geographically large state such as Queensland, where services are centralised in major centres. This phenomenon has been described previously, where older age was associated with reduced willingness to travel for specialist care.^22^ The increased severity of AS pre-procedurally in regional and remote patients further supports potential delays in diagnosis, workup and treatment for these groups. Because our study relies on a procedural registry, these upstream disparities cannot be measured, and patients who required TAVR but were unable to access it due to rurality are not captured.

Younger age may also explain the lower STS score observed as rurality increased, as age is a major component of the score. Alternatively, there may be lower patient or clinician risk tolerance in rural areas, a hypothesis that we cannot substantiate with previous research and requires further investigation. First Nations people were also underrepresented in this dataset, accounting for 0.6% of the cohort compared with 5.2% of Queensland’s population according to Australian Bureau of Statistics estimates.^16^ Differences in procedural characteristics, including transfemoral access, balloon-expanding valve use and cerebral protection device use, most likely reflect preferences of the treating hospitals.

Of the predictors of 12-month mortality that we observed, previous AF or flutter and clinical frailty score were not statistically different between rurality groups. Transfemoral access route was high in all groups and its variation across groups followed no consistent rurality related pattern that would explain mortality differences. The higher STS score yet lower chronic lung disease in urban patients may counterbalance any mortality disparities. More broadly, the absence of outcome disparities likely reflects the centralised TAVR model of Queensland, whereby all patients receive their TAVR in hospitals that comply with the same regulatory standards. Encouragingly, it also suggests equitable access to post-procedural care, which in Queensland involves a combination of tertiary specialist review, local cardiology input, general practitioner follow-up, cardiac rehabilitation, and allied health support^23^; although, the extent to which these models are accessed is not captured in ACOR. This finding applies only to patients who reached TAVR, and the 12-month follow-up period may be insufficient to detect longer-term divergence.

The most recent report of ACOR TAVR data indicates national 12-month all-cause mortality after TAVR to be 5.6%, which is slightly higher than that of the urban (5.0%), regional (4.9%) and remote (4.8%) groups in Queensland reported in this study.^24^ These 12-month mortality outcomes were also superior to real-world TAVR registries internationally, where reported one-year mortality has generally ranged from approximately 9.6% to 17.0%, although comparisons are limited by differences in treatment era, patient risk profile and procedural practice.^25–28^ Furthermore, the null rurality effect for TAVR outcomes is consistent with previous literature, and is now confirmed in a larger sample with multivariable time-to-event analysis.^29,30^

This analysis demonstrated that the median length of hospital stay was one day longer for patients in regional and remote Queensland compared to those in urban areas. This was statistically significant; however, it is unclear whether this indicates delayed recovery or procedural complications necessitating prolonged hospitalisation. We hypothesise that this may have arisen due to travel logistics for regional and remote patients, although if attributable to avoidable hospital days, this difference may have cost implications, with one Australian estimate valuing a hospital bed day at $AUD2871.^31^ The marginally greater improvements in valve area and mean gradient among regional and remote patients may reflect more severe baseline AS. In contrast, the small differences in LVEF improvement are likely clinically trivial and may reflect inter-observer variability. The higher paravalvular regurgitation at 30 days in urban patients, while not upheld at post-procedural and 12-month echocardiograms, may be a result of the greater use of self-expanding valves in this group, which are known to have higher rates than balloon expanding valves.^32^

Findings of the current study must be considered in line with inherent strengths and limitations. To our knowledge, this is the largest international study examining TAVR outcomes in the context of rurality and the first Australian study to capture remote patients. Additionally, it utilises data from a high-quality national registry with mandated reporting, and sensitivity analyses confirmed the stability of results. A key limitation in the current dataset is that there is some disparity between MMM classification and Australian postcodes. For example, a single postcode may contain several MMM regions leading to potential misclassification of rurality and unmeasured bias. To mitigate this, we assigned the least rural MMM classification in this scenario, based on the assumption that most residents lived in the less rural area. We do acknowledge that this could bias our results towards the null. Furthermore, as discussed above, the use of a TAVR registry means only patients who received a TAVR were captured and disparities in diagnosis or referral were not measured. Finally, the retrospective design leaves room for unmeasured confounding.

In conclusion, this study has confirmed that rurality alone should not be assumed to confer higher procedural risk and helps guide future research. The centralised TAVR delivery in Queensland appears to be equitable based on the results from this procedure-based registry, although future research needs to assess access pathways rather than procedural outcomes. Future studies linking echocardiography or diagnostic data to TAVR data could better assess whether rural patients with severe AS are underdiagnosed, referred later or less likely to receive TAVR and if their overall AS outcomes are disparate. Prospective cohort studies are also required to reduce the risk of confounding. Furthermore, ArcGIS could be used to more accurately assign patients MMM classification to reduce the risk of bias related to incorrect MMM classification and perform geospatial modelling of outcomes. Finally, the follow-up period could be extended to assess longer-term outcome data.

## Conclusion

In conclusion, we have confirmed that rurality is not an independent predictor of 12-month TAVR mortality in a large, multicentre study utilising data from the high-quality ACOR. We also descriptively confirmed that rurality does not impact other clinical or echocardiographic outcomes. This is despite rural patients presenting with younger age, greater comorbidity burden, more severe AS but lower STS scores. These results pave the way for future research focusing on ascertaining if access disparities exist at the diagnostic, referral or workup stage of the AS pathway, which may in turn inform the future planning and development of non-metropolitan TAVR services.

## Author contributions

**Patrick Cameron:** conceptualisation, methodology, formal analysis, investigation, data curation, writing-original draft, writing-review and editing, visualisation, project coordination. **Joseph Moxon:** Methodology, validation, formal analysis, writing-review and editing, visualisation, supervision. **Anthony Brazzale:** conceptualisation, supervision, writing-review and editing. **Greg Starmer:** conceptualisation, supervision, writing-review and editing.

Patrick Cameron and Joseph Moxon directly accessed and verified the underlying participant-level data.

All authors had full access to the data and accept responsibility to submit.

## Data sharing

Individual participant data will not be made available to others. The data were provided by ACOR under a data-use agreement and the conditions of the ethics approval for this study, which do not permit release of participant-level data outside the research team. Readers are welcome to contact the corresponding author with any questions about the study.

## Abbreviations

ACOR: Australian Cardiac Outcomes Registry
AF: atrial fibrillation
AR: aortic regurgitation
AS: aortic stenosis
ASGS-RA: Australian Statistical Geography Standard–Remoteness Areas
AV: aortic valve
AVA: aortic valve area
BAV: balloon aortic valvuloplasty
BMI: body mass index
CI: confidence interval
CKD: chronic kidney disease
eGFR: estimated glomerular filtration rate
FUP: follow-up
GP: general practitioner
HF: heart failure
HR: hazard ratio
HREC: Human Research Ethics Committee
IECD: implanted cardiac electrical device
IQR: interquartile range
LOS: length of stay
LVEF: left ventricular ejection fraction
MACCE: major adverse cardiac and cerebrovascular events
MI: myocardial infarction
MMM: Modified Monash Model
NYHA: New York Heart Association
PCI: percutaneous coronary intervention
PPM: permanent pacemaker
SAVR: surgical aortic valve replacement
SPSS: Statistical Package for the Social Sciences
STS: Society of Thoracic Surgeons
TAVR: transcatheter aortic valve replacement
TAVI: transcatheter aortic valve implantation
TIA: transient ischaemic attack

## Acknowledgements

Thank you to the ACOR staff who supplied the data. There was no funding source for this study.

During the preparation of this work the authors used Claude Sonnet 5.5 (Anthropic) to draft R code for the Kaplan–Meier curve (Figure 1) and to calculate Benjamini–Hochberg adjusted p values (q values) for Tables 1 and 2.

## References

1. Osnabrugge RLJ, Mylotte D, Head SJ, et al. Aortic Stenosis in the Elderly: Disease Prevalence and Number of Candidates for Transcatheter Aortic Valve Replacement: A Meta-Analysis and Modeling Study. J Am Coll Cardiol. 2013;62(11):1002–1012. doi:10.1016/j.jacc.2013.05.015

2. Nkomo VT, Gardin JM, Skelton TN, Gottdiener JS, Scott CG, Enriquez-Sarano M. Burden of valvular heart diseases: a population-based study. The Lancet. 2006;368(9540):1005–1011. doi:10.1016/S0140-6736(06)69208-8

3. Otto CM. Timing of aortic valve surgery. Heart. 2000;84(2):211–218. doi:10.1136/heart.84.2.211

4. Leon MB, Smith CR, Mack M, et al. Transcatheter Aortic-Valve Implantation for Aortic Stenosis in Patients Who Cannot Undergo Surgery. N Engl J Med. 2010;363(17):1597–1607. doi:10.1056/NEJMoa1008232

5. Bennetts J, Sinhal A, Walters D, et al. 2021 CSANZ and ANZSCTS Position Statement on the Operator and Institutional Requirements for a Transcatheter Aortic Valve Implantation (TAVI) Program in Australia. Heart Lung Circ. 2021;30(12):1811–1818. doi:10.1016/j.hlc.2021.07.017

6. Clune SJ, Blackford J, Murphy M. Management of the acute cardiac patient in the Australian rural setting: a 12 month retrospective study. Aust Crit Care Off J Confed Aust Crit Care Nurses. 2014;27(1):11–16. doi:10.1016/j.aucc.2013.03.002

7. Rural and remote health. Australian Institute of Health and Welfare. November 20, 2025. Accessed June 23, 2026. https://www.aihw.gov.au/reports/rural-remote-australians/rural-and-remote-health

8. Clark RA, Coffee N, Turner D, et al. Application of Geographic Modeling Techniques to Quantify Spatial Access to Health Services Before and After an Acute Cardiac Event. Circulation. 2012;125(16):2006–2014. doi:10.1161/CIRCULATIONAHA.111.083394

9. Roule V, Rebouh I, Lemaitre A, et al. Impact of wait times on late postprocedural mortality after successful transcatheter aortic valve replacement. Sci Rep. 2022;12(1):5967. doi:10.1038/s41598-022-09995-z

10. Wijeysundera HC, Wong WWL, Bennell MC, et al. Impact of Wait Times on the Effectiveness of Transcatheter Aortic Valve Replacement in Severe Aortic Valve Disease: A Discrete Event Simulation Model. Can J Cardiol. 2014;30(10):1162–1169. doi:10.1016/j.cjca.2014.03.009

11. Elbaz-Greener G, Yarranton B, Qiu F, et al. Association Between Wait Time for Transcatheter Aortic Valve Replacement and Early Postprocedural Outcomes. J Am Heart Assoc. 2019;8(1):e010407. doi:10.1161/JAHA.118.010407

12. Arnold SV, Afilalo J, Spertus JA, et al. Prediction of Poor Outcome After Transcatheter Aortic Valve Replacement. JACC. 2016;68(17):1868–1877. doi:10.1016/j.jacc.2016.07.762

13. Ludman PF, Moat N, de Belder MA, et al. Transcatheter aortic valve implantation in the United Kingdom: temporal trends, predictors of outcome, and 6-year follow-up: a report from the UK Transcatheter Aortic Valve Implantation (TAVI) Registry, 2007 to 2012. Circulation. 2015;131(13):1181–1190. doi:10.1161/CIRCULATIONAHA.114.013947

14. Field PE, Franklin RC, Barker RN, Ring I, Leggat PA. Cardiac rehabilitation services for people in rural and remote areas: an integrative literature review. Rural Remote Health. 2018;18(4). doi:10.22605/RRH4738

15. Zahn R, Gerckens U, Linke A, et al. Predictors of One-Year Mortality After Transcatheter Aortic Valve Implantation for Severe Symptomatic Aortic Stenosis. Am J Cardiol. 2013;112(2):272–279. doi:10.1016/j.amjcard.2013.03.024

16. Region summary: Queensland. Australian Bureau of Statistics. Accessed October 22, 2024. https://dbr.abs.gov.au/region.html?lyr=steCrgn=3.

17. Australasian Cardiac Outcomes Registry. ACOR. Accessed April 25, 2026. https://acor.net.au/.

18. Modified Monash Model. Australian Government Department of Health, Disability and Ageing. April 10, 2025. Accessed June 10, 2026. https://www.health.gov.au/topics/rural-health-workforce/classifications/mmm?language=en

19. Postal Areas (2021) by Modified Monash Model (2023). Data.gov.au. November 6, 2025. Accessed September 16, 2026. https://data.gov.au/data/dataset/postal-areas-2021-by-modified-monash-model-2023

20. Versace VL, Skinner TC, Bourke L, Harvey P, Barnett T. National analysis of the Modified Monash Model, population distribution and a socio-economic index to inform rural health workforce planning. Aust J Rural Health. 2021;29(5):801–810. doi:10.1111/ajr.12805

21. Heart, stroke and vascular disease: Australian facts. Australian Institute of Health and Welfare. June 17, 2024. Accessed October 22, 2024. https://www.aihw.gov.au/reports/heart-stroke-vascular-diseases/hsvd-facts/contents/about

22. Bühn S, Holstiege J, Pieper D. Are patients willing to accept longer travel times to decrease their risk associated with surgical procedures? A systematic review. BMC Public Health. 2020;20(1):253. doi:10.1186/s12889-020-8333-5

23. Going home after a Transcatheter Aortic Valve Procedure. The Prince Charles Hospital. March 2015. Updated March 2018. Accessed September 18, 2026. https://metronorth.health.qld.gov.au/tpch/wp-content/uploads/sites/3/2017/08/Going-Home-after-TAVI.pdf.

24. Sinhal A, Bennetts J, Bhindi R, et al. Australian Cardiac Outcomes Registry of Transcatheter Aortic Valve Implantation: Report and Update of Transcatheter Aortic Valve Implantation in Australia. Heart Lung Circ. 2025;34(5):472–484. doi:10.1016/j.hlc.2025.02.107

25. Rudolph TK, Herrmann E, Bon D, et al. Comparison of contemporary transcatheter heart valve prostheses: data from the German Aortic Valve Registry (GARY). Clin Res Cardiol. 2024;113(1):75–85. doi:10.1007/s00392-023-02242-z

26. Overtchouk P, Guedeney P, Rouanet S, et al. Long-Term Mortality and Early Valve Dysfunction According to Anticoagulation Use. JACC. 2019;73(1):13–21. doi:10.1016/j.jacc.2018.08.1045

27. Yamamoto M, Watanabe Y, Tada N, et al. Transcatheter aortic valve replacement outcomes in Japan: Optimized CathEter vAlvular iNtervention (OCEAN) Japanese multicenter registry. Cardiovasc Revasc Med. 2019;20(10):843–851. doi:10.1016/j.carrev.2018.11.024

28. Carroll JD, Mack MJ, Vemulapalli S, et al. STS-ACC TVT Registry of Transcatheter Aortic Valve Replacement. JACC. 2020;76(21):2492–2516. doi:10.1016/j.jacc.2020.09.595

29. Paleri S, Tham JLM, Jin D, et al. Transcatheter aortic valve implantation for severe aortic stenosis in the Australian regional population. Aust J Rural Health. 2019;27(3):3. doi:10.1111/ajr.12508

30. Fogelson B, Baljepally R, Heidel E, et al. Rural versus urban outcomes following transcatheter aortic valve implantation: The importance of the heart team. Cardiovasc Revasc Med. 2024;62:3–8. doi:10.1016/j.carrev.2023.12.005

31. Partington A, Karnon J. Revealing the Value of a Bed Day: The Health Economists’ Study Group Summer 2025 Meeting. In: 2025. Accessed September 10, 2026. https://hesg.org.uk/meetings/summer-2025-office-of-health-economics/

32. Van Belle E, Vincent F, Labreuche J, et al. Balloon-Expandable Versus Self-Expanding Transcatheter Aortic Valve Replacement: A Propensity-Matched Comparison From the FRANCE-TAVI Registry. Circulation. 2020;141(4):243–259. doi:10.1161/CIRCULATIONAHA.119.043785

